# Association of Low-Dose Mood Stabilizers With Treatment Retention in Unipolar Depression: A Retrospective Cohort Study

**DOI:** 10.1101/2025.06.04.25328948

**Authors:** Hideyo Sugahara, Shoji Tokunaga

**Affiliations:** Sugahara Tenjin Clinic, GG Solar Building, Tenjin 3-4-9 Chuo-ku, Fukuoka-city, Fukuoka 810-0001, Japan; Clinical Research Support Center, Ehime University Hospital, 454 Shitsukawa, Toon City, Ehime Prefecture, 791-0295, Japan

## Abstract

**Importance:** Mood stabilizers are typically reserved for bipolar disorders, yet their role in treating unipolar depression remains underexplored despite clinical need and theoretical rationale.

**Objective:** To evaluate the association between low-dose mood stabilizer use and 6-month treatment retention in patients with unipolar and bipolar depression.

**Design, Setting, and Participants:** This retrospective cohort study analyzed medical records from a single outpatient psychiatric clinic in Japan between 2016 and 2022, including 300 patients with unipolar or bipolar depression initiating pharmacotherapy (100 patients per year).

**Exposures:** Prescription of mood stabilizers (e.g., lithium, valproate), antidepressants, or both.

**Main Outcomes and Measures:** The primary outcome was 6-month treatment retention, used as a proxy for pharmacotherapy adherence, reflecting long-term drug tolerability and real-world clinical effectiveness. Retention rates were compared across treatment types using Kaplan–Meier and Cox regression analyses adjusted for demographic and clinical variables.

**Results:** Mood stabilizer prescriptions increased from 27% in 2016 to 89% in 2022. Patients with unipolar depression treated with mood stabilizers exhibited significantly higher 6-month retention compared to those on antidepressants alone (adjusted HR: 0.44; 95% CI, 0.24–0.80; p <.01). Side effects were mild and manageable, with high continuation rates observed for low-dose regimens.

**Conclusions and Relevance:** Low-dose mood stabilizers, particularly lithium and valproate, were associated with significantly higher treatment retention rates in unipolar depression compared to antidepressants alone. These findings suggest the potential role of mood stabilizers in enhancing treatment adherence and highlight the need for confirmatory prospective studies to evaluate their comparative effectiveness.

**Key Points:** *Question:* Can low-dose mood stabilizers improve 6-month treatment retention among patients with unipolar depression?

*Findings:* In this retrospective cohort study of 185 patients with unipolar depression, treatment with low-dose lithium or valproate was associated with significantly higher 6-month treatment retention compared to antidepressants alone (adjusted HR: 0.44; 95% CI, 0.24–0.80; p <.01).

*Meaning:* Low-dose mood stabilizers may improve treatment adherence in unipolar depression, warranting reconsideration of their role in treatment guidelines.

---

Despite pharmacological advances, unipolar depression remains a leading cause of global disability,^1,2^ with challenges in treatment adherence^3,4^ and relapse prevention.^5,6^ Although antidepressants dominate treatment guidelines,^7,8^ their effectiveness in ensuring adherence^9,10^ and preventing suicide^11,12^ remains suboptimal, prompting exploration of alternatives like mood stabilizers.^13^ Previous studies have demonstrated lithium’s effectiveness in reducing suicide risk^14,15^ and managing treatment-resistant depression,^16,17^ yet its use in unipolar depression remains limited.^18-21^ The DSM-III’s dichotomous classification^22,23^ may have contributed to underrecognition of what is called bipolar spectrum^24-26^ presentations and limited the therapeutic application of mood stabilizers. Furthermore, the rapid expansion of antidepressant medications^27,28^—often supported by short-term trials—has overshadowed evidence for alternative approaches.

This study aimed to evaluate the association between mood stabilizer use and six-month treatment adherence in patients with unipolar and bipolar depression, addressing the observed increase in their prescription and potential clinical benefits.

## Methods

### Study Design

This retrospective cohort study was designed to evaluate the impact of mood stabilizers on depressive disorders in light of recent increases in their prescription. Medical records of 300 patients with mood disorders (100 patients per year) from 2016, 2019, and 2022 were reviewed. Key variables assessed included patient demographics, prescribed mood stabilizers and antidepressants, and the frequency of hospital visits within the initial six months of treatment. Hospital visits and medication adherence are essential for evaluating drug effectiveness.^29-31^ Therefore, we used hospital visit and prescription records as the drug efficacy indicator.^32-35^ The sample size was calculated to achieve 80% power at a two-sided significance level of 5%, assuming a six-month continuation rate of 35% in 2016 and 50% in 2022, requiring a minimum of 97 participants per year. The study was approved by the Ethics Committee of Tohoku University and complied with the STROBE guidelines.

### Participants

A total of 100 consecutive patients per year were included, beginning on or after April 1 of each respective year. Inclusion criteria were a diagnosis of unipolar or bipolar depression and initiation of treatment at our clinic. Exclusion criteria encompassed visits unrelated to treatment (e.g., family consultations, second opinions), treatment for non-depressive psychiatric or medical conditions, cases not requiring continuous pharmacotherapy (e.g., mild depression, anxiety, or insomnia), diagnostic-only visits, and treatment refusal (Figure 1). The starting date was chosen to minimize seasonal variation.

**Figure 1.**
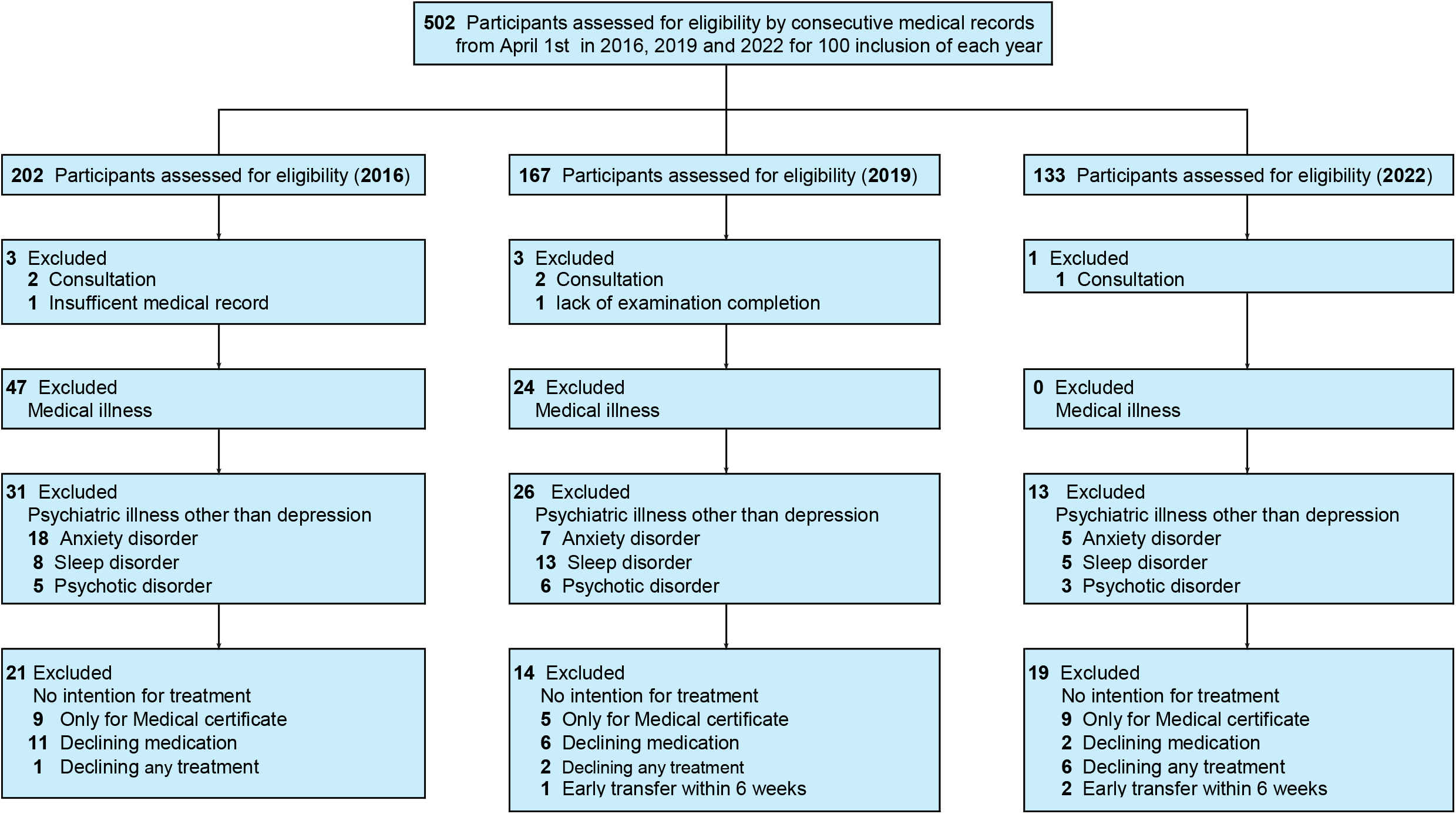
Flowchart of participant inclusion. Participants were included based on the order of their initial visit date. Exclusion criteria included patients with medical illness or psychiatric conditions other than depression, those without the intention for standard treatment at the clinic, and inappropriate cases such as family consultations. Other reasons for exclusion included an insufficient medical record (2016) and a lack of examination completion at the clinic (2019).

Initial assessments included a comprehensive health questionnaire, including the Center for Epidemiologic Studies Depression Scale (CES-D),^36^ a 30-minute nursing interview, and a 40–60-minute physician consultation. Treatment adhered to standard guidelines^7,8,37^ recommending medication continuation for 6–9 months after symptom improvement in single episodes and longer for recurrent cases. Although DSM-5 criteria^38^ were used, diagnoses of bipolar depression incorporated additional considerations such as subthreshold manic symptoms, affective temperament, and family history, consistent with a bipolar spectrum model. ^39-41^

In 2016, mood stabilizers were predominantly prescribed for bipolar disorder, but their use evolved over time. By 2022, they were also prescribed for conditions such as atypical depression or exhaustion-type depression with suspected mood instability. Off-label use was disclosed to patients, and informed consent was obtained. Patients were informed about the approved indications of lithium and valproate and advised that prescriptions beyond these indications were off-label. Informed consent was documented, and verbal checks were routinely conducted during follow-ups. Suicidal ideation data were extracted from binary responses on intake questionnaires. Patients with persistent suicidal ideation or risk behaviors were treated with risperidone or other antipsychotics as needed. Anxiolytics and hypnotics were prescribed for acute or intermittent symptoms.

### Investigation

Descriptive statistics were computed for baseline variables. It was assumed all patients received mood stabilizers and/or antidepressants. The mean maximum dose and duration of medication use over six months were calculated. Six-month treatment retention rates were analyzed using the Kaplan-Meier method and stratified by age, sex, diagnosis (unipolar or bipolar), suicidal ideation, and CES-D score. Between-year differences were evaluated using Cox regression adjusted for these variables. Due to missing suicidal ideation responses from three patients in 2022, this analysis was conducted on 297 (203 unipolar and 94 bipolar) participants. When evaluating antidepressant and mood stabilizer effects based on treatment adherence, hospital visit duration from the initial consultation cannot reflect the effects of medications introduced later. The group that began concomitant use of the two kinds of drugs after the initial visit had already elapsed some time by the time combination therapy was initiated, so these cases need to be excluded when comparing the administration periods with each monotherapy group. The analysis excluded 18 patients with unipolar depression receiving both medications for this reason. The final analysis included 185 unipolar depression cases. Among unipolar cases, adherence was compared across groups receiving antidepressants, mood stabilizers, or both.

Factors influencing six-month retention and reasons for early discontinuation were identified from medical records by two independent nurses, utilizing predefined criteria based on patient-reported outcomes and physician assessments. Disagreements were resolved through consensus. Categorical reasons were aggregated and ranked by frequency. This approach supplements hospital visit records and prescription data, providing comprehensive understanding of medication adherence behaviors.

### Statistical Analysis

ANOVA was used to assess differences in means, while Kaplan-Meier and Cox regression analyses were employed to assess hospital visit retention. All statistical analyses were conducted using EZR^42^ (ver. 1.68), a graphical user interface for R, with statistical significance defined as p <.05. Potential biases inherent to the retrospective design were addressed by adjusting for demographic and clinical variables in the Cox regression models.

## Results

### Patients

Across the three cohorts (2016, 2019, 2022), a total of 300 patients were included. The mean age was 33.1 years (SD 12.6), and 70% were female. The proportion of patients diagnosed with unipolar depression ranged from 59% to 78% across the years (Table 1). Mood stabilizer prescriptions increased from 27% in 2016 to 54% in 2019 and 89% in 2022 (Table2).

**Table 1.** Demographic Characteristics of the Participants.

| Year | 2016 | 2019 | 2022 |
| --- | --- | --- | --- |
| N | 100 | 100 | 100 |
| Mean age (years) | 36.5 (13.5) | 32.9 (13.2) | 30.1 (10.0) |
| Age range (years) | 15~75 | 13~75 | 14~59 |
| Women/Men | 63/37 | 77/23 | 69/31 |
| Major depressive disorder (n) | 78 | 59 | 67 |
| Bipolar disorder (n) | 22 | 41 | 33 |
| Suicidal ideation (n) | 33 | 39 | 64 (unknown: 3) |
| Mean CES-D* score | 35.4 (11.9) | 35.7 (10.3) | 38.5(9.4) |
| Survey Period** |  |  |  |
| Survey Start Date | April 1 | April 1 | April 1 |
| Final Day of Survey | October 19 | September 10 | August 10 |

Parentheses show the standard deviation.
\*CES-D: The Center for Epidemiologic Studies Depression Scale
\*\*The chart review was conducted consecutively in the order of the initial consultation dates.

**Table 2.** Mood Stabilizer and Antidepressant Prescriptions.

| Year | 2016 | 2019 | 2022 |
| --- | --- | --- | --- |
| *Mood Stabilizer* |  |  |  |
| Li only (n) | 20 | 37 | 70 |
| Mean maximum dose (mg) | 315.0(134.8) | 216.2(130.2) | 251.4(122.8) |
| Mean dosing period (weeks) | 14.5(10.9) | 17.1(11.2) | 15.4(11.0) |
| VAP only (n) | 6 | 12 | 7 |
| Mean maximum dose (mg) | 300.0(154.9) | 300.0(165.2) | 171.4(125.4) |
| Mean dosing period (weeks) | 19.3(11.8) | 12.5(11.8) | 13.2(10.5) |
| Combination (Li/VAP) (n) | 0 | 4 | 12 |
| Mean maximum dose (mg) | — | 175.0(82.9)/175.0(43.3) | 283.3(103.0)/183.3(71.8) |
| Mean dosing period (weeks) | — | 7.0(8.5)/11.0(3.8) | 18.3(11.3)/16.5(8.0) |
| Lamotrigine (n) | 1 | 1 | 0 |
| Net participants with any prescription (n) | 27 | 54 | 89 |
| *Antidepressant (n)* |  |  |  |
| Sulpiride | 47 | 41 | 56 |
| Sertraline | 20 | 14 | 7 |
| Trazodone | 5 | 1 | 28 |
| Escitalopram | 11 | 6 | 3 |
| Mirtazapine | 9 | 5 | 6 |
| Paroxetine | 3 | 11 | 0 |
| Amitriptyline | 2 | 4 | 7 |
| Others* | 13 | 9 | 1 |
| Net participants with any prescription | 84 | 80 | 76 |

Parentheses show the standard deviation.
Li: lithium carbonate; VAP: sodium valproate
\*Other antidepressants include fluvoxamine (8), duloxetine (6), mianserin (6), maprotiline (2), and nortriptyline (1).

### Outcome

#### Retention by Year

Kaplan–Meier survival analysis of 6-month treatment adherence rates over 3 years (n=297), adjusted for age, sex, illness, presence of suicidal ideation and CES-D score, showed an increase over time (Figure 2). Cox proportional hazards analysis, adjusting for the same variables, indicated a significant year effect on hospital visits (hazard ratio [HR]: 0.81; 95% confidence interval [CI]: 0.67–0.99; p=.03), suggesting increased attendance period over time.

**Figure 2.**
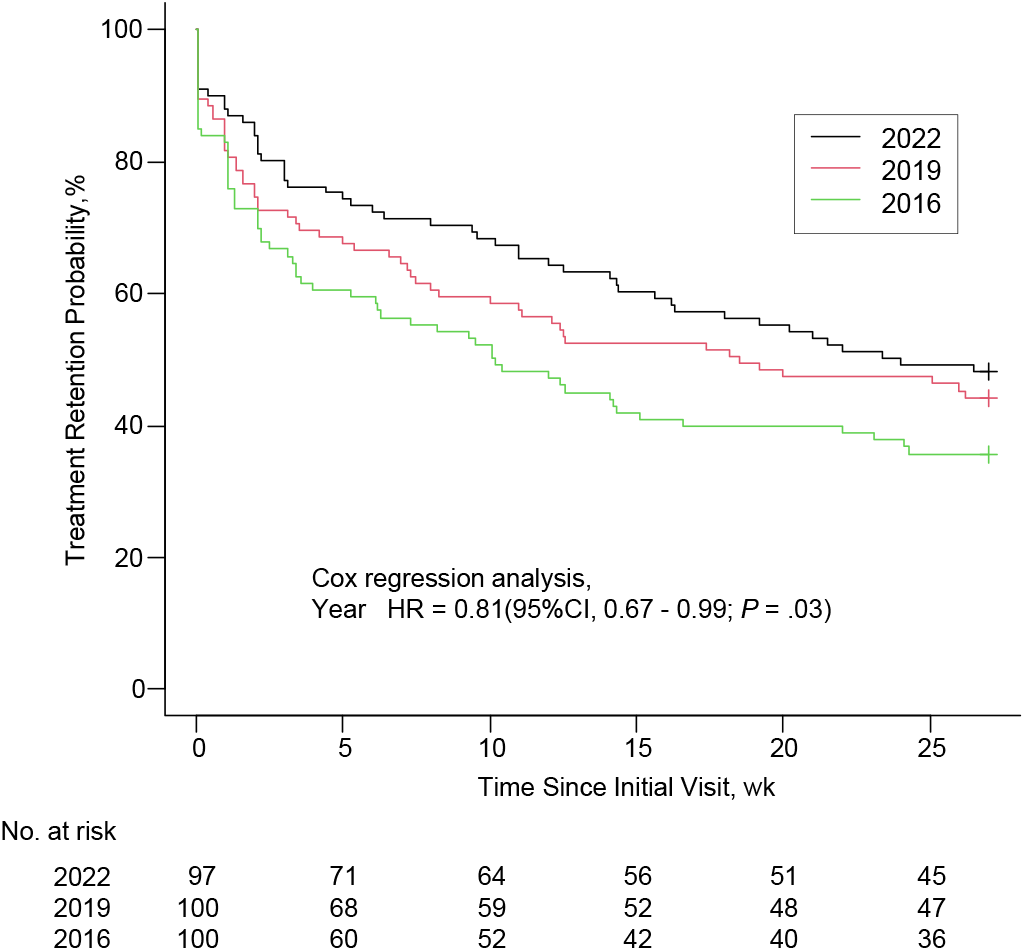
Kaplan–Meier and Cox regression analyses comparing treatment retention across 2016, 2019 and 2022. The analysis depicts 6-month retention rates among all participants for each year. Cox regression analysis was adjusted for age, sex, illness, the presence of suicidal ideation (SI), and Center for Epidemiologic Studies Depression Scale (CES-D) score. CI, confidence interval

#### Retention by Treatment Group

Patients with unipolar depression (n=185) receiving mood stabilizers demonstrated significantly higher 6-month retention compared to those on antidepressants alone (adjusted HR: 0.44; 95% CI, 0.24–0.80; p <.01). These findings are visually represented in Figure 3, highlighting the divergence in retention rates over time.

**Figure 3.**
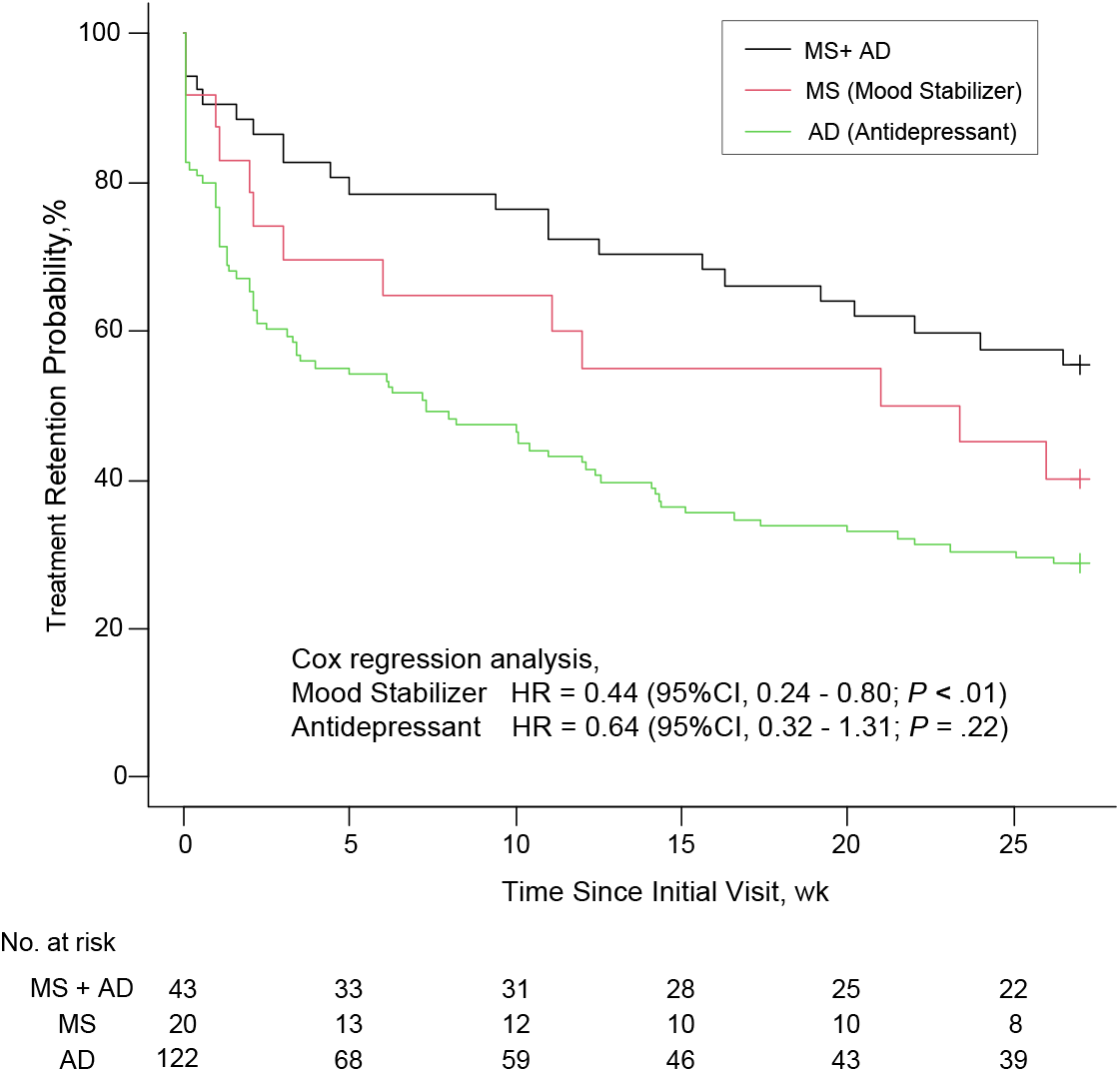
Kaplan–Meier and Cox regression analyses of treatment retention in unipolar depression subgroup (n=185) The analysis compares treatment adherence among participants receiving mood stabilizers, antidepressants, or both. Cox regression analysis was adjusted for age, sex, the presence of suicidal ideation (SI), Center for Epidemiologic Studies Depression Scale (CES-D) score, and year. In the Cox regression model, mood stabilizer (MS) and antidepressant (AD) were represented as binary variables, with 0 indicating not prescribed and 1 indicating prescribed. For all participants diagnosed with unipolar depression (n=185), hazard ratios for MS and AD were calculated, with adjustment for these covariates. CI, confidence interval

#### Assumed Reasons for Treatment Adherence

Patients who continued treatment frequently reported subjective improvement or insufficient improvement, while early discontinuation was often attributed to lack of perceived benefit or social factors. Reasons for continuation or dropout were consistently categorized across cohorts.

#### Safety

Routinely monitored side effects were mild and manageable in most cases, including gastrointestinal discomfort and tremors in patients taking lithium and somnolence in those on valproate. Dosages were typically low (lithium: 100–600 mg/day; valproate: 100–600 mg/day), contributing to good tolerability.

## Discussion

This retrospective cohort study suggests that mood stabilizers may be associated with improved treatment retention for both bipolar and unipolar depression. Prescription rates increased from 27% in 2016 to 89% in 2022, with improved 6-month retention rates. Patients with unipolar depression receiving mood stabilizers showed higher retention rates, with an adjusted hazard ratio of 0.44. These findings suggest mood stabilizers may enhance treatment adherence even without a bipolar diagnosis.^21,43,44^ The expanded use of mood stabilizers was driven by the clinician’ careful monitoring of patient outcomes, which provided compelling evidence for their effectiveness. The diagnostic distribution showed a 2:1 ratio of unipolar to bipolar depression, differing from U.S. DSM-IV’s 5:1 ratio.^38^ Over 45% of patients reported suicidal ideation at baseline, rising to 65% by 2022. Antidepressants may pose risks by inducing manic symptoms, highlighting mood stabilizers’ importance for suicide prevention.^45,46^ While lithium has proven anti-suicidal properties^14,15^ and efficacy in treatment-resistant depression,^16,17^ its use in unipolar depression remains limited in Western countries where antidepressants dominate.^18-21^ The increasing prescription of mood stabilizers suggests a shift in clinical practice with global implications. The tolerability of low-dose mood stabilizers suggests utility in diverse settings including their potential to reduce relapse rates and improve patients’ quality of life. Future studies should explore whether these benefits extend to other clinical outcomes, such as functional recovery and suicide prevention, particularly in populations with high baseline suicidal ideation.

Pharmaceutical market dynamics and antidepressant proliferation^27,28^—often approved after brief trials—have overshadowed mood stabilizers. Few new mood stabilizers have been introduced, and lithium remains underprescribed, despite favorable outcomes and minimal side effects at low doses. In this study, adverse events were mild and manageable, supporting broader application of mood stabilizers. Social factors including the COVID-19 pandemic and healthcare changes may have influenced results.^47,48^ The increase in the proportion of suicidal ideation among participants in this study in 2022 is assumed to reflect this impact. However, the clinic environment remained stable. Patient reports of improvement were common reasons for continued visits, suggesting perceived treatment benefit.

Low-dose mood stabilizers offer a promising strategy to enhance treatment adherence in unipolar depression, along with clinical benefits and potential ethical advantages, such as reducing inappropriate antidepressant monotherapy in patients with latent bipolarity. These findings underscore the need for prospective randomized controlled trials comparing adherence and efficacy across diverse populations, including antidepressant monotherapy and combination therapy with antidepressants and mood stabilizers, in order to validate their effectiveness and inform revisions to international treatment guidelines.

### Limitations

While this study provides preliminary evidence supporting the use of low-dose mood stabilizers in unipolar depression, several limitations must be addressed. The retrospective design precludes definitive conclusions about causality, highlighting the need for prospective randomized controlled trials to confirm these findings. Moreover, the use of hospital visits and prescription records as proxies for treatment adherence may not always perfectly capture patient behavior or treatment motivation. Additionally, as the study was conducted at a single clinic in Japan, the generalizability of the results to other healthcare systems or cultural contexts remains uncertain. Future multicenter studies across diverse populations are essential to validate these results. Furthermore, the findings underscore the need to reconsider the role of mood stabilizers in international treatment guidelines, particularly in regions where their use remains limited. Comparative studies examining the efficacy of mood stabilizers in different healthcare settings could provide valuable insights into their broader applicability.

## Conclusions

This study demonstrates that mood stabilizers, particularly lithium and valproate at low doses, are associated with improved treatment retention and manageable side effects in patients with depression including those diagnosed with unipolar depression. These findings highlight the potential role of mood stabilizers to enhance long-term treatment adherence and support the need for future research to inform guideline revisions.

## Data Availability

All relevant data are provided within the manuscript and its tables and figures.

## Acknowledgements

The authors are grateful to Akane Yoshida, Haruka Taniguchi, Honoka Matsunobu, Chie Koga, Fumine Matsubara, and Mei Shirouzu for their contributions to data acquisition and methodological discussions related to this research.

